# Tafazzin regulates neutrophil maturation and inflammatory response

**DOI:** 10.1101/2024.06.05.24307331

**Authors:** Przemysław Zakrzewski, Christopher M. Rice, Kathryn Fleming, Drinalda Cela, Sarah J. Groves, Fernando Ponce, Willem Gibbs, Kiran Roberts, Tobias Pike, Douglas Strathdee, Eve Anderson, Angela H. Nobbs, Ashley Toye, Colin Steward, Borko Amulic

**Author notes:** Contributed equally (Przemysław Zakrzewski, Christopher M. Rice).

## Abstract

Barth syndrome (BTHS) is a rare genetic disease caused by mutations in the *TAFAZZIN* gene. It is characterized by neutropenia, cardiomyopathy and skeletal myopathy. Neutropenia in BTHS is associated with life-threatening infections, yet there is little understanding of the molecular and physiological causes of this phenomenon. We combined bone marrow analysis, CRISPR/Cas9 genome editing in hematopoietic stem cells and functional characterization of circulating BTHS patient neutrophils to investigate the role of *TAFAZZIN* in neutrophils and their progenitors. We demonstrate a partial cell intrinsic differentiation defect, along with a dysregulated neutrophil inflammatory response in BTHS, including elevated formation of neutrophil extracellular traps (NETs) in response to calcium flux. Developmental and functional alterations in BTHS neutrophils are underpinned by perturbations in the unfolded protein response (UPR) signaling pathway, suggesting potential therapeutic avenues for targeting BTHS neutropenia.

## INTRODUCTION

Barth syndrome (BTHS) is a rare X-linked genetic disease characterized principally by dilated cardiomyopathy, skeletal myopathy, and neutropenia [1–3]. Mutations in the *TAFAZZIN* gene, encoding a mitochondrial lipid transacylase, are the primary cause of BTHS [4]. Tafazzin is essential for remodeling and maturation of cardiolipin (CL), a major phospholipid of the mitochondrial inner membrane, which has a crucial role in maintaining mitochondrial structure and function [2, 5]. *TAFAZZIN* mutations lead to an accumulation of an intermediate CL species, monolysocardiolipin (MLCL), impairing mitochondrial metabolism and contributing to defects in cardiomyocytes and skeletal muscle, which have abundant mitochondria and are dependent on efficient oxidative respiration [6]. Apart from regulating ATP generation, CL has been implicated in signaling [7, 8], control of apoptosis [8, 9], generation of mitochondrial reactive oxygen species (ROS) [9], and regulation of calcium homeostasis [10]. Furthermore, mitochondria are important regulators of inflammation, and it has been suggested that dysregulated inflammatory response is a component of BTHS [11], although there is a major lack of knowledge on the innate immune response in patients. Neutropenia and neutrophil-mediated inflammation are possible targets for therapeutic interventions, to improve patient wellbeing and survival.

Neutropenia is detected in approximately 84% of BTHS patients and shows various patterns [1, 3, 12], including intermittent and unpredictable, chronic or severe, or truly cyclical [12]. Neutrophils kill microbes by phagocytosis, generation of ROS, release of antimicrobial proteins stored in secretory granules, and formation of neutrophil extracellular traps (NETs), which consist of externalized chromatin decorated with antimicrobial proteins [13]. NETs can trap microbes and prevent their dissemination but are also sensed by other innate and adaptive immune cells, leading to proinflammatory cytokine production and propagation of inflammation [14]. Importantly, neutropenia in BTHS is associated with various bacterial infections, which are the second leading cause of death in these patients [1].

Despite the prominence of neutropenia in BTHS patients, there is little understanding of the molecular and physiological causes of this phenomenon [12]. Neutropenia can result from defects in neutrophil production in the bone marrow, elevated apoptosis or from enhanced removal of activated neutrophils from the circulatory system [15–17].

Limited investigation of bone marrow in BTHS led to conflicting reports: hypocellularity and reduced myeloid maturation were detected in some BTHS patients without a clear block in neutrophil maturation [12, 18], and short-term colony assays using hematopoietic stem cells (HSCs) purified from patient blood failed to detect any defects in neutrophil differentiation *in vitro* [18, 19]. One study described elevated annexin V binding in circulating BTHS neutrophils, without increased apoptosis or phagocytic clearance by macrophages [19]. On the other hand, studies using shRNA-mediated *TAFAZZIN* knockdown in myeloid cell lines demonstrated increased caspase-3 activation, release of cytochrome c from mitochondria, and accelerated apoptosis in response to tafazzin deficiency [20]. There is therefore no consensus on the mechanism of neutropenia in BTHS patients.

In mice, tafazzin deficiency causes embryonic lethality in C57BL/6 mice [21, 22], although other genetic backgrounds are viable. Immortalization of murine embryonic *Tafazzin*-KO myeloid progenitors demonstrated normal differentiation into mature neutrophils, normal mitochondrial electron transport chain assembly, and equivalent functional responses to wild-type (WT) cells [23]. The study did, however, detect slightly enhanced susceptibility to apoptosis upon Bcl-2 inhibition and subtle perturbations in expression of unfolded protein response (UPR) genes, raising the possibility that ER-mediated stress may affect viability of BTHS cells [23].

To address the gap in understanding of BTHS neutropenia, we combined patient bone marrow analysis with our recently developed *ex vivo* neutrophil differentiation system [24] to investigate the role of *TAFAZZIN* in neutrophil development from HSCs. We also performed a comprehensive analysis of neutrophil phenotype and function in BTHS patients, including proteomic analysis. Our work reveals a partial block in neutrophil differentiation and maturation in BTHS, accompanied by elevated intercellular calcium, hyperdegranulation, and evidence of enhanced NET formation. We detect alterations in the UPR pathway and propose that this may be a potential cause of the observed defects in BTHS neutrophils.

## RESULTS

### Partial block in bone marrow neutrophil maturation in BTHS

We examined neutrophil morphology in bone marrow aspirates of 5 BTHS patients (4-24 years old; all treated with G-CSF). The samples had similar features and were characterized by normocellularity, normal megakaryopoiesis and erythropoiesis, and unaffected eosinophil lineage (Fig. 1A, S1A). As expected with G-CSF treatment, all samples exhibited marked ‘left shift’ i.e. an elevated number of granulocyte precursors – promyelocytes and myelocytes. However, there was a notable decrease in the number of metamyelocytes and mature neutrophils compared to values observed in healthy individuals; this was reflected in substantially reduced myeloid to erythroid ratio (Fig. 1B). These findings demonstrate a defect in neutrophil maturation in BTHS, as previously suggested [12, 18].

**Fig. 1.**
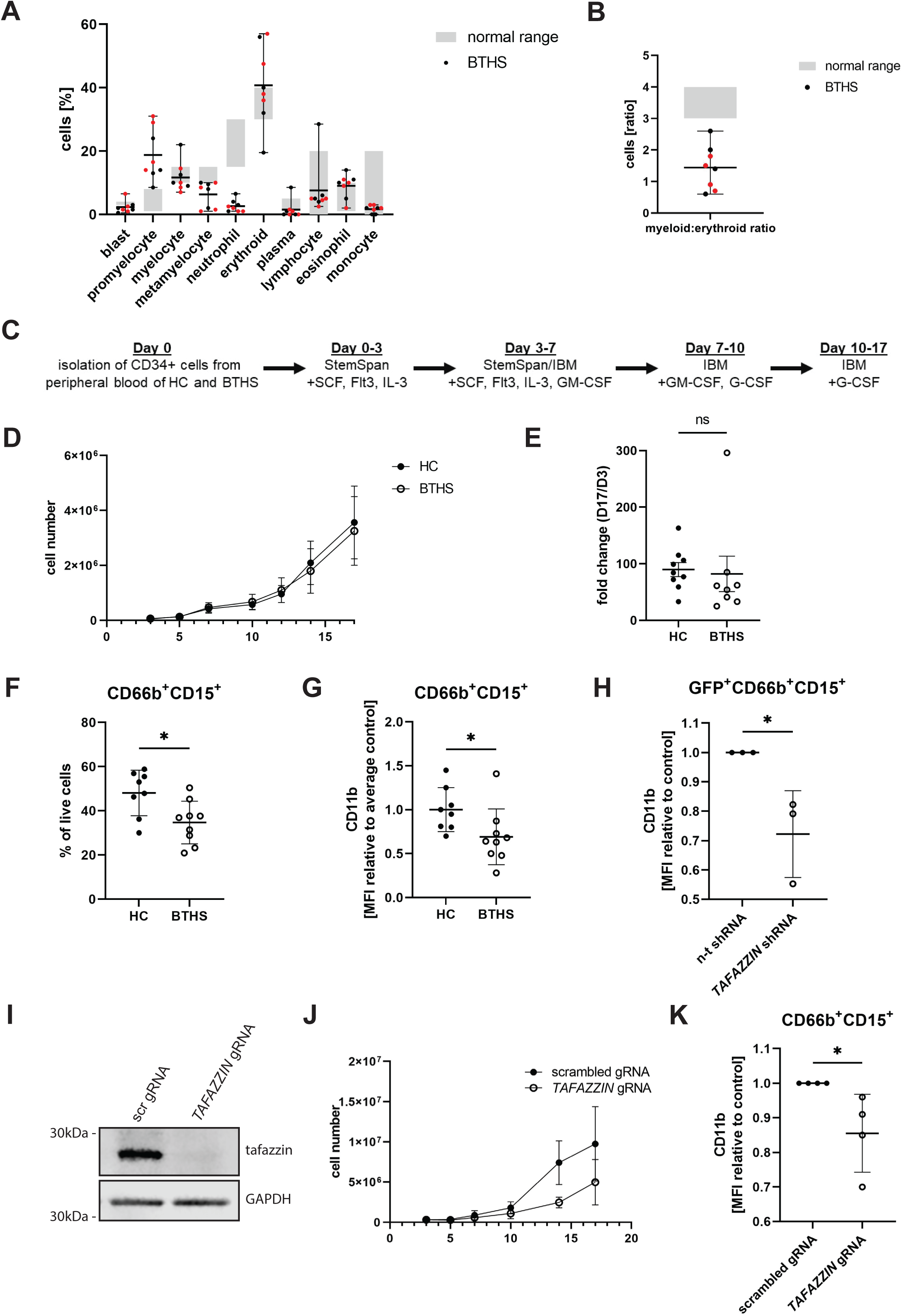
Tafazzin regulates neutrophil development. **A.** Wright-Giemsa staining of 8 bone marrow aspirates from BTHS patients. The normal range (grey) represents values for healthy children. Samples from the same patient, obtained at different ages, are marked in red. **B.** Myeloid:erythroid ratio for samples analysed in A. **C.** Simplified overview of culture protocol for deriving neutrophils from HSC. **D.** Growth curve of *ex vivo* HSC-derived neutrophil, n = 9 (HC), 8 (BTHS). **E.** Fold change of total cell count of HSC-derived neutrophils from day 3 to day 17 of differentiation, n = 9 (HC), 8 (BTHS); ns – not significant. **F.** Percent of CD66b^+^CD15^+^ neutrophils in HSC-derived cells at the end of differentiation, n = 9 (HC), 8 (BTHS); * P ≤ 0.05. **G.** CD11b surface expression of CD66b^+^CD15^+^ HSC-derived neutrophils at the end of differentiation, relative to averaged CD11b expression of control cells on a same day, n = 9 (HC), 8 (BTHS); * P ≤ 0.05. **H.** CD11b surface expression of control or tafazzin shRNA-treated CD66b^+^CD15^+^GFP^+^ HSC-derived neutrophils at the end of differentiation, relative to n-t shRNA control, n = 9 (HC), 8 (BTHS); * P ≤ 0.05. **I.** Representative tafazzin Western blot of CRISPR/Cas9-edited HSC-derived neutrophils at the end of differentiation. **J.** Growth curve of CRISPR/Cas9-edited HSC-derived neutrophils, n = 3 (experimental repeats). **K.** CD11b surface expression of CD66b^+^CD15^+^ CRISPR/Cas9-edited HSC-derived neutrophils at the end of differentiation, relative to scrambled gRNA control, n = 4; * P ≤ 0.05.

### Impaired *ex vivo* terminal differentiation and maturation of BTHS neutrophils

To date, there have been no studies of maturation of tafazzin deficient neutrophils using primary human progenitors. Our laboratory recently optimized a protocol to differentiate human neutrophils from CD34^+^ HSCs isolated from a small amount of peripheral blood (Fig. 1C, adapted from [24]). This protocol yields approximately 50% mature neutrophils, defined as CD66b^+^CD15^+^ cells. We used this system to compare neutrophil differentiation from CD34^+^ progenitors of healthy control (HC) and BTHS patients, recruited from the NHS National Barth Syndrome Service at Bristol Royal Hospital for Children. We did not observe any difference in total cell number over 17 days of cell differentiation (Fig. 1D) or in proliferation ratio of BTHS and HC cells (fold change from day 3: HC = 90±37 vs. BTHS = 82±88); (Fig 1E). Moreover, there was no increase in apoptosis or necrosis in a subset of BTHS cultured neutrophils (Fig. S1B) or any evident morphological changes (Fig. S1C). However, flow cytometry (Fig. S1D) uncovered an average 13.4% drop in the number of differentiated CD66b^+^CD15^+^ neutrophils (HC = 48.08±10.30% vs. BTHS = 34.69±9.68%; p = 0.0145); (Fig. 1F) and a 31% decrease in relative expression of the maturity marker CD11b in BTHS samples (HC = 1.00±0.25 vs. BTHS = 0.69±0.32; p = 0.0425); (Fig. 1G). Importantly, this reduction was also reflected by an average 12.7% drop in the number of mature CD66b^+^CD15^+^CD11b^hi^ neutrophils at the end of differentiation (HC = 37.08±5.80% vs. BTHS = 24.40±7.26%; p = 0.0013) (Fig. S1E).

To confirm a cell-intrinsic role for tafazzin in control of neutrophil differentiation, we performed shRNA knockdown experiments. First, to choose an optimal shRNA construct, we used the myeloid cell line PLB-985 to screen 3 vectors encoding *TAFAZZIN*-targeting shRNA. The most consistent results were obtained with shRNA 3, where we observed reductions in both expression of tafazzin and expression of CD11b in differentiated cells (Fig. S1F). Next, we isolated CD34^+^ HSC from healthy donors and treated them with control non-targeting or *TAFAZZIN*-targeting shRNA, on day 3 of differentiation (corresponding to non-committed progenitors), which led to an approximately 45% reduction in tafazzin expression (Fig. S1G). Similarly to BTHS patient cultured neutrophils, *TAFAZZIN* shRNA progenitors differentiated less efficiently than controls, as evidenced by a 28% reduction in CD11b expression at the end of differentiation (Fig. 1H).

Finally, to test the effect of complete tafazzin deficiency, we used CRISPR/Cas9 to target exon 3 in CD34^+^ HSCs (day 3/non-committed progenitors, n = 3), with a scrambled gRNA serving as control. Western analysis confirmed complete tafazzin knockout at day 17 of differentiation (Fig. 1I). Importantly, complete tafazzin deficiency resulted in an approximately 50% reduction in cell proliferation, compared to scrambled control, when comparing fold expansion between days 3 and 17 (scrambled gRNA = 33±15 vs. *TAFAZZIN* gRNA = 17±9, p = 0.1394) (Fig. 1J, S1H). Moreover, similarly to shRNA-treated neutrophils, tafazzin knockout cells exhibited a ∼15% decrease in the expression of differentiation marker CD11b (Fig. 1K). In summary, experiments using HSC-derived neutrophils demonstrate that tafazzin has a cell autonomous role in regulating neutrophil differentiation and maturation from early progenitors and confirm a partial block in granulopoiesis in BTHS patients.

### Immature status of circulating BTHS neutrophils

We next examined circulating neutrophils in BTHS patients (n = 15, 4/15 not receiving G-CSF therapy) and healthy controls (n = 15, all untreated). Flow cytometric immunophenotyping of neutrophil surface markers in whole blood, using previously described gating strategies [25], revealed significantly reduced levels of the maturity marker CD10 (HC = 4036±2343 vs. BTHS = 2168±1969; p = 0.0253) and a trend towards reduced expression of CD16 (FcYRIIIB) (HC = 20222±10030 vs. BTHS = 14037±9303; p = 0.0908); (Fig. 2A). We did not observe any differences in CD101 and CD62L between HC and BTHS patients (Fig. S2B). Both CD10 and CD16 expression were normal in G-CSF untreated BTHS patients (n=4, ‘non-neutropenic patients’); (Fig. S2A). It must be stressed that these patients do not suffer from documented neutropenia, suggesting that their hematopoietic compartment is not affected. G-CSF mobilizes immature neutrophils from the bone marrow, resulting in accumulation of circulating neutrophils with reduced CD10 and CD16 [26], making it difficult to conclude whether the observed immaturity is due to G-CSF treatment or an intrinsic feature of BTHS neutrophils.

**Fig. 2.**
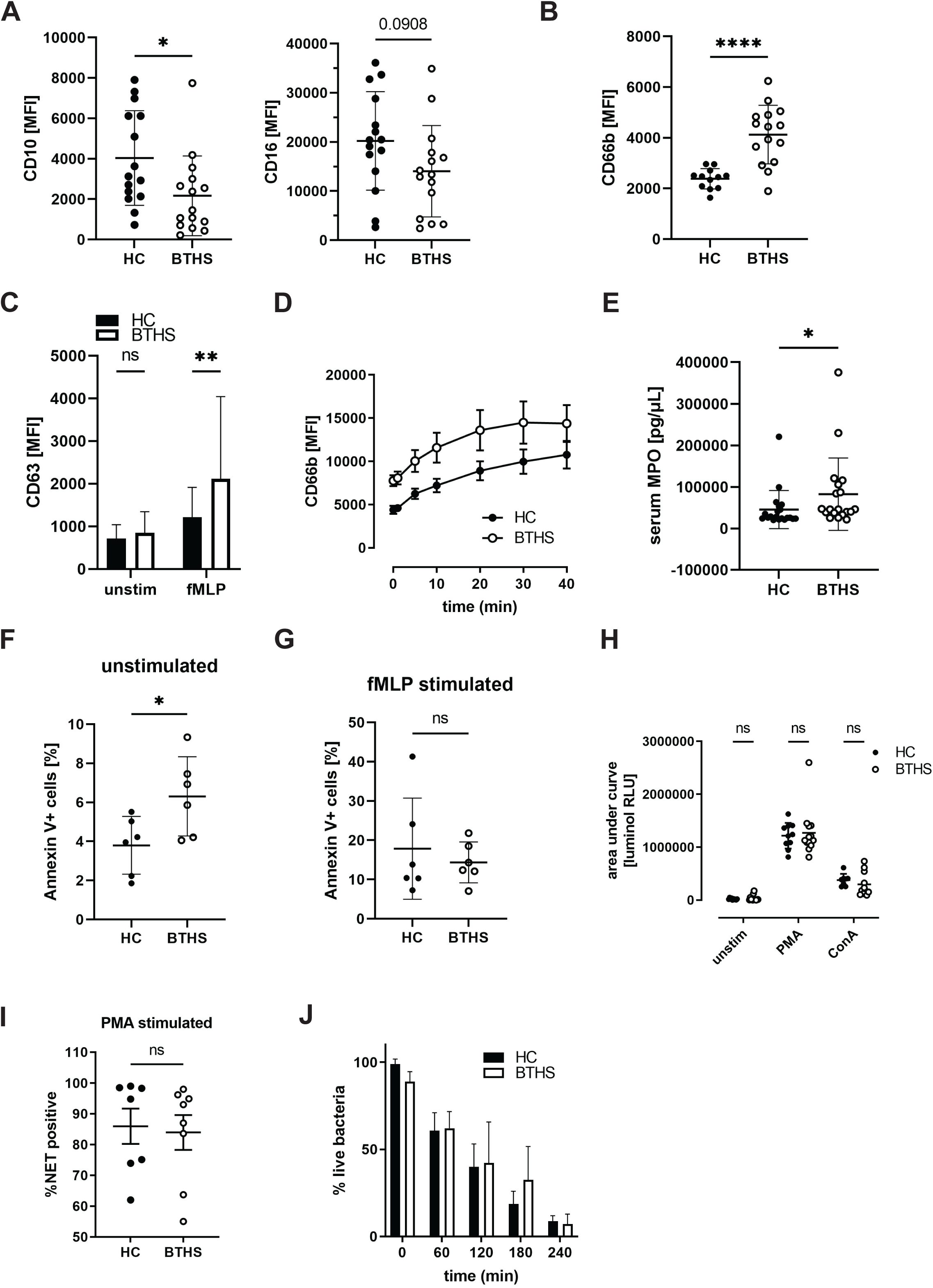
Characterization of peripheral blood neutrophils from BTHS patients. **A.** Surface expression of CD10 (left) and CD16 (right) in circulating neutrophils, n = 15 (HC, BTHS); * P ≤ 0.05. **B.** Surface expression of CD66b in circulating neutrophils, n = 12 (HC), 15 (BTHS); *** P ≤ 0.001. **C.** Surface expression of CD63 in unstimulated (unstim) and fMLP-stimulated (300 nM) isolated neutrophils, n (unstim) = 29 (HC, BTHS), n (fMLP) = 31 (HC, BTHS); ns – not significant, ** P ≤ 0.01. **D.** Time-course of CD66b surface expression in isolated neutrophils stimulated with *Streptococcus pyogenes*, MOI = 100, n = 9 (HC), 7 (BTHS). **E.** MPO concentration in plasma, n = 20 (HC), 19 (BTHS); * P ≤ 0.05. **F.** Percentage of annexin V+ unstimulated neutrophils, n = 6 (HC, BTHS); * P ≤ 0.05 **G.** Percentage of annexin V+ neutrophils, after stimulation with 300 nM fMLP, n = 6 (HC, BTHS); ns – not significant. **H.** Area under the curve (AUC) quantification of ROS, detected by luminol, n (unstim, PMA) = 10 (HC), 12 (BTHS), n (ConA) = 8 (HC), 10 (BTHS); ns – not significant. **I.** NET release in response to 50 nM PMA, n = 7 (HC), 8 (BTHS); ns – not significant. **J.** Quantification of viable *Streptococcus pyogenes* bacteria relative to “serum only” control, after incubation with isolated neutrophils, n = 4 (HC, BTHS).

### BTHS neutrophils exhibit elevated degranulation and competent bacterial killing

We next used flow cytometry to examine neutrophil surface markers associated with activation. Interestingly, the secondary granule marker CD66b expression was significantly elevated on patient neutrophils (HC = 2385±401 vs. BTHS = 4125±1156; p < 0.0001), suggesting increased *in vivo* exocytosis compared to HC (Fig. 2B). This finding was independent of G-CSF therapy (Fig. S2C). To examine degranulation in more detail, including exocytosis of primary granules, we isolated neutrophils using negative selection and quantified exposure of the primary granule marker CD63, with or without stimulation with the bacterial peptide *N*-formylmethionine-leucyl-phenylalanine (fMLP). There was no significant difference in CD63 exposure in naïve neutrophils. Upon fMLP stimulation, however, BTHS neutrophils (n=31) demonstrated more than 1.7-fold increase in activation compared to HC (n=29); (HC = 1213±700 vs. BTHS = 2115±1927; p = 0.0032); (Fig. 2C). To test if elevated degranulation occurs in response to live bacteria, we stimulated purified neutrophils with *Streptococcus pyogenes* and quantified exposure of CD66b over 40 minutes of co-incubation (Fig. 2D). We found elevated surface levels of the secondary granule marker in naïve BTHS neutrophils, confirming our *in vivo* findings, as well as an approximately 2-fold increase upon bacterial stimulation compared to HC. Finally, we quantified plasma abundance of MPO, a primary granule protein, and found elevated circulating MPO levels in BTHS patients compared to healthy control plasma samples (Fig. 2E). In summary, neutrophils from BTHS patients are more prone to degranulation of primary and secondary granules.

A previous report demonstrated elevated rates of phosphatidyl serine (PS) externalization in BTHS neutrophils, although these levels were lower than those typically associated with neutrophil apoptosis [19]. We stained a portion of our neutrophils with annexin V to quantify PS surface exposure. As previously reported, we observed increased PS exposure in naïve neutrophils (HC = 3.80±1.48% vs. BTHS = 6.31±2.02%; p = 0.0344); (Fig. 2F). To test whether elevated PS exposure in BTHS neutrophils could result from increased degranulation, we demonstrated that fMLP stimulation increases annexin V positivity approximately 3-fold both for HC and BTHS neutrophils compared to unstimulated cells (HC = 17.86±12.88% vs. BTHS = 14.36±5.19%; not significant); (Fig. 2G).

Secondary granules contain multiple cytoadhesive molecules and augmented degranulation might result in excessive endothelial interactions, leading to sequestration of neutrophils in vascular beds. We therefore analyzed spleen and liver histological sections from WT and *Tafazzin*-KO mice (Fig. S2D). We did not observe any difference in the number of neutrophils in the vasculature and parenchyma of these tissues, indicating normal levels of margination in mice under steady state conditions. Interestingly, in contrast to what is observed in BTHS patients, *Tafazzin*-KO mice, on FVB (sensitive to the Friend leukemia virus) genetic background, had equivalent neutrophil counts to those of WT mice, suggesting no defects in hematopoiesis (Fig. S2E).

Next, we investigated whether neutrophils from BTHS patients show other functional alterations. Using a luminol based assay, we quantified ROS production in response to phorbol 12-myristate 13-acetate (PMA) and concanavalin A (con A) and found no significant differences compared to HC (Fig. 2H). Moreover, we found no difference in NETs formation with the strong soluble inducer PMA (Fig. 2I). Moreover, BTHS neutrophils phagocytosed pHrodo^TM^ *Streptococcus pyogenes* at similar rates to HC neutrophils (Fig. S2F). Finally, neutrophils from BTHS patients (n=4) and HC (n=4) demonstrated equivalent killing rates of opsonized *S. pyogenes* bacteria (Fig. 2J). In summary, BTHS neutrophils display enhanced secondary and primary degranulation, normal PMA-induced NETosis and maintain effective killing of a Gram-positive pathogen.

### Mitochondrial function is not affected in BTHS patient neutrophils

Mitochondria in BTHS patient neutrophils have not previously been studied. We found increased mitochondrial abundance in patients relative to control, quantified with MitoTracker dye (HC = 1.00±0.22 vs. BTHS = 1.66±0.52; p = 0.001); (Fig. 3A). Interestingly, BTHS patient neutrophils had reduced mitochondrial membrane potential, as evidenced by reduction of tetramethylrhodamine ethyl ester (TMRE) compared to HC neutrophils (HC = 31±19% TMRE low cells vs. BTHS = 47±25% TMRE low cells; p = 0.0349); (Fig. 3B), suggesting that the increased abundance may be a compensatory mechanism for impaired activity. To test this hypothesis, we used Seahorse metabolic flux analysis to test respiratory chain activity in isolated BTHS primary neutrophils (n = 8, 4/8 treated with G-CSF) and HC. We found no significant differences in mitochondrial ATP production, basal respiration, maximal respiration, or spare respiratory capacity (Fig. 3C), arguing against gross functional impairment.

**Fig. 3.**
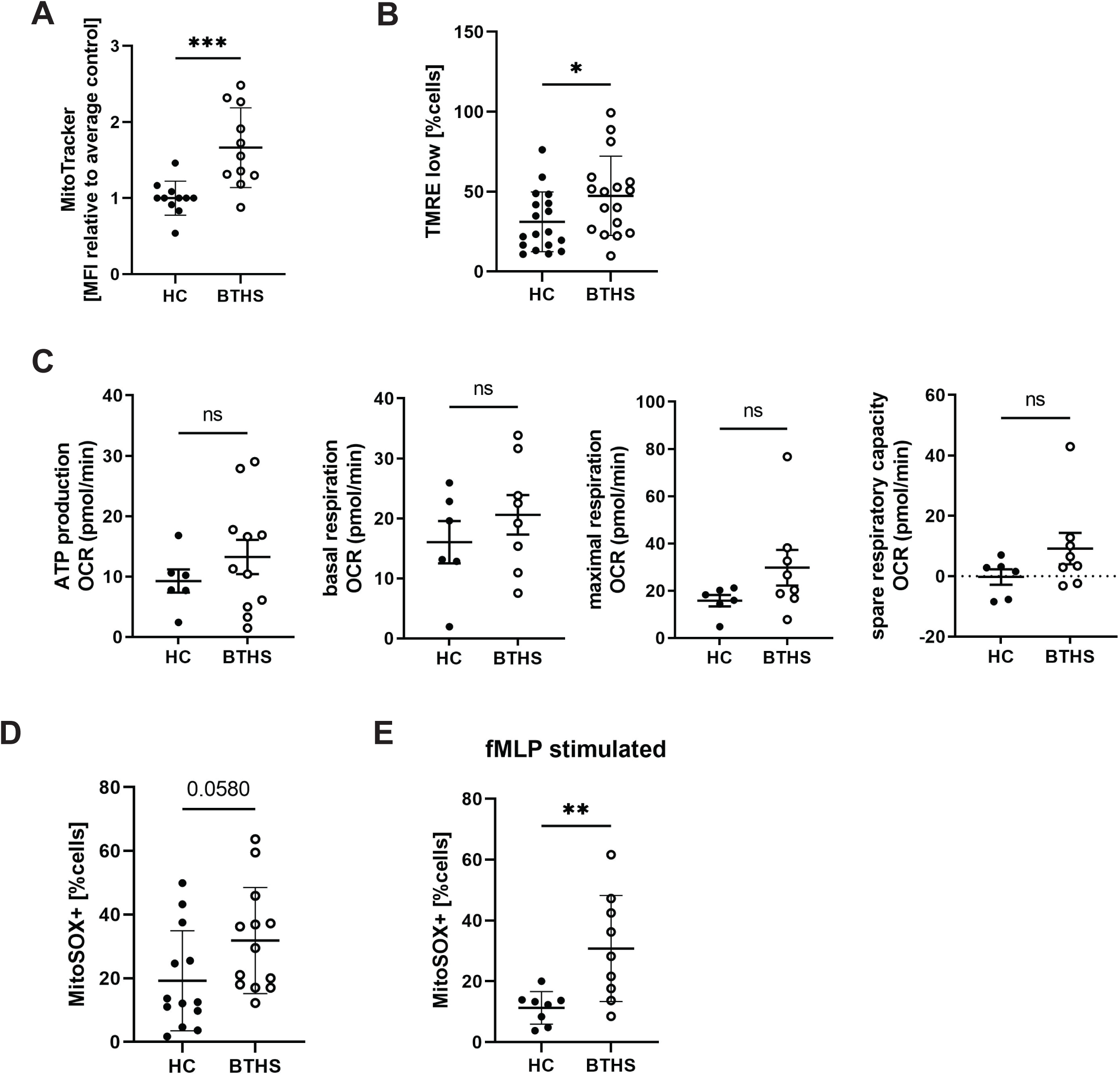
Mitochondrial perturbations in BTHS patient circulating neutrophils. **A.** Average MitoTracker median fluorescence of isolated BTHS patient circulating neutrophils normalized to HC (on the same day), n = 11 (HC, BTHS); *** P ≤ 0.001. **B.** Average percentage of neutrophils with low TMRE signal, n = 18 (HC), 17 (BTHS); * P ≤ 0.05. **C.** Average rates of ATP production, basal respiration, maximal respiration, and spare respiratory capacity, measured by Seahorse metabolic flux analyzer, n (ATP) = 6 (HC), 11 (BTHS), n (basal, maximal, spare respiration) = 6 (HC), 8 (BTHS); ns – not significant. **D.** Percentage of MitoSOX-positive neutrophils n = 13 (HC, BTHS). **E.** Percentage of MitoSOX-positive neutrophils, after stimulation with 300 nM fMLP, n = 8 (HC), 9 (BTHS); ** P ≤ 0.01.

Finally, as several studies reported elevated mitochondrial ROS (mtROS) in BTHS cardiomyocytes [2, 27], we quantified mtROS in BTHS neutrophils using a MitoSOX flow cytometry assay. The percentage of MitoSOX-positive cells was elevated in BTHS samples, both in homeostatic conditions (HC = 19.20±15.72 vs. BTHS = 31.85±16.66; p = 0.058); (Fig. 3D) and after stimulation with fMLP (HC = 11.27±5.35% vs. BTHS = 30.79±17.46%; p = 0.0084); (Fig. 3E), although this difference was not observed when plotting total MitoSOX MFI (Fig. S3A-B). Moreover, we did not detect any mtROS differences in BTHS HSC-derived neutrophils in a limited number of samples (Fig. S3C-D). In conclusion, mitochondria in GCSF-treated BTHS patients are characterized by elevated abundance, reduced membrane potential and elevated mtROS. However, we failed to detect significant impairment of overall mitochondrial functionality.

### Increased calcium-induced NETosis in BTHS neutrophils

Excessive mtROS can affect calcium homeostasis, which in turn can regulate neutrophil activation [28]. As we observed increased mtROS in BTHS neutrophils, we decided to quantify intracellular calcium level in these neutrophils using X-Rhod-1, a cell permeant dye that exhibits increased fluorescence after binding Ca^2+^. We found a >2-fold increase in intracellular calcium level of circulating BTHS cells, compared to HC neutrophils (HC = 1.00±0.08 vs. BTHS = 2.33±1.26; p = 0.0025); (Fig. 4A). To determine whether this change is caused by G-CSF therapy or a cell-intrinsic effect of tafazzin deficiency, we analyzed calcium levels in HSC-derived neutrophils and observed an almost 3-fold increase in X-Rhod-1 signal in BTHS stem cell-derived neutrophils, compared to HC (HC = 2570±671 vs. BTHS = 5662±1160; p = 0.0162); (Fig. 4B). Calcium is an essential activator of neutrophil inflammatory responses [29], prompting us to ask whether tafazzin deficient neutrophils have altered functional responses. To test this, we utilized the calcium ionophore A23187, which induces calcium influx across the plasma membrane and strongly activates neutrophils [30]. As expected, A23187 induced strong degranulation of primary granules, but we did not observe any significant differences in CD63 surface exposure between control and tafazzin CRISPR/Cas9 knockout HSC-derived neutrophils (Fig. 4C).

**Fig. 4.**
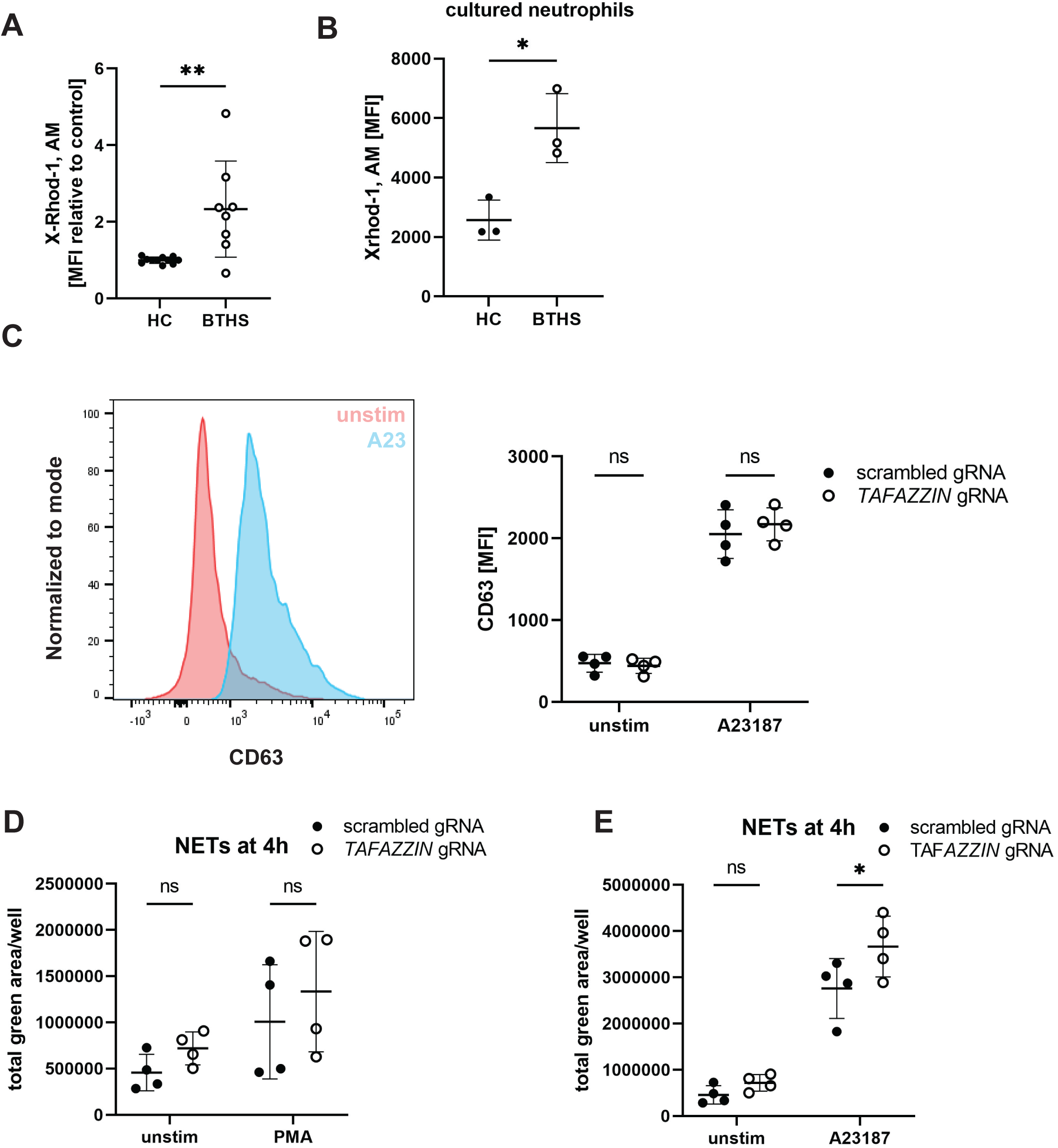
Calcium homeostasis in BTHS neutrophils. **A.** Average median fluorescence of calcium dye X-Rhod-1, normalized to HC (on the same day), n = 11 (HC), 8 (BTHS); ** P ≤ 0.01. **B.** Quantification of average median fluorescence of X-Rhod-1 in HSC-derived BTHS neutrophils (D17), n = 3 (HC, BTHS); * P ≤ 0.05. **C.** An example of histogram depicting CD63 surface expression in HSC-derived neutrophils before and after stimulation with A23187 and surface expression of CD63 in unstimulated (unstim) and A23187-stimulated (2.5 μM) CRISPR/Cas9-edited HSC-derived neutrophils, n (unstim) = 4, n (A23) = 4; ns – not significant. **D.** Quantification of NET release by CRISPR/Cas9-edited HSC-derived neutrophils before or after stimulation with PMA (50nM) at 4 h post-stimulation, n = 4; ns – not significant. **E.** Calculation of NET release by CRISPR/Cas9-edited HSC-derived neutrophils before or after stimulation with A23187 (10 μM) at 4 h post-stimulation, n = 4; ns – no significant, * P ≤ 0.05.

We next investigated NET formation in tafazzin knockout HSC-derived neutrophils in response to calcium-independent (PMA) and calcium-dependent stimuli (A23187), using a live cell imaging assay. There was no difference in chromatin release in response to PMA (Fig. 4D). On the other hand, A23187-induced NETosis was significantly increased in tafazzin knockout cells compared to control cells (Fig. 4E). In conclusion, both circulating and stem cell derived BTHS neutrophils exhibit elevated intracellular calcium concentration, which promotes higher rates of NET formation in response to calcium ionophore.

### Elevated UPR signaling in BTHS patients

To better understand developmental changes in BTHS patient neutrophils, we performed tandem mass tag (TMT) proteomic analysis of circulating neutrophils from patients (n = 4 G-CSF-treated and n = 1 untreated, non-neutropenic patient), as well as healthy controls (n = 5). We detected a total of 4056 proteins (Fig. S5A), including 306 proteins with significantly altered expression (p < 0.05), 91 of which had >2-fold difference. Ingenuity Pathway Analysis (IPA) revealed multiple significantly up– and down-regulated pathways in BTHS neutrophils (Fig. 5A). Notably, the most strongly enriched pathway in BTHS was oxidative phosphorylation (Fig. S5B), closely followed by fatty acid oxidation (Fig. S5C), consistent with elevated mitochondrial load, as detected with MitoTracker (Fig. 3C). Intriguingly, the second most enriched pathway in BTHS neutrophils was the unfolded protein response (UPR), which was previously implicated in neutrophil dysregulation in *Tafazzin*-KO mice [23]. Specifically, we observed an upregulation of eight UPR proteins (Fig. 5B) and four proteins associated with mitochondrial UPR (mt-UPR, Fig. 5C). These changes appeared to be GCSF– and neutropenia-dependent, as the only untreated, non-neutropenic patient in our study consistently exhibited less pronounced alteration (sample *BTHS5*). Of note, changes in the expression of mt-UPR proteins were more uniform among patients (Fig. S5D).

**Fig. 5.**
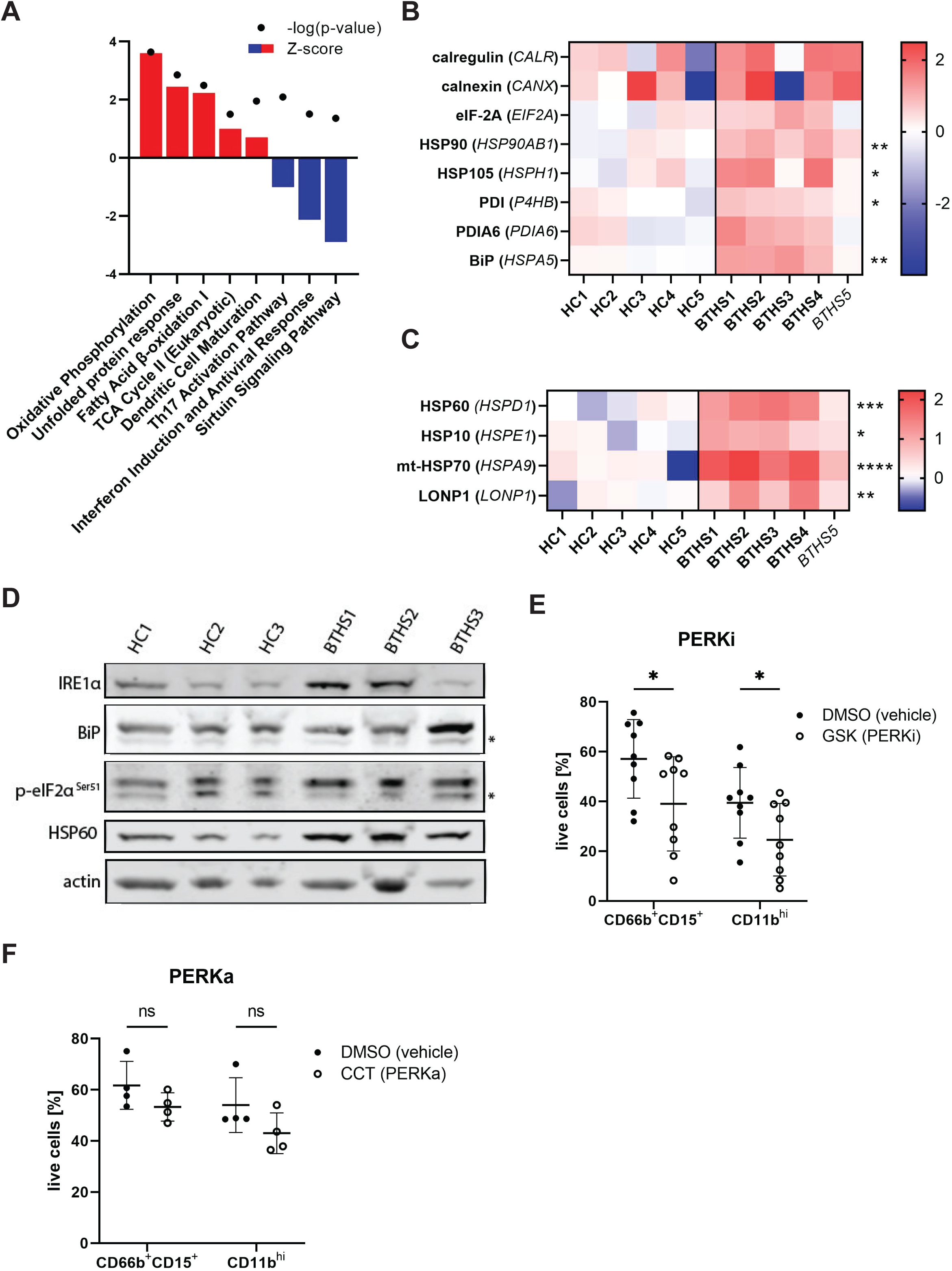
Elevated UPR signaling in BTHS neutrophils. **A.** Plot showing significantly upregulated (positive Z-score, red) and downregulated (negative Z-score, blue) IPA canonical pathways in circulating neutrophils from BTHS patients. **B.** Heat map depicting upregulated UPR-related proteins in circulating BTHS neutrophils, displayed as log2 fold change over HC, * P ≤ 0.05, ** P ≤ 0.01. **C.** Heat map depicting upregulated mitochondrial UPR-related proteins in circulating BTHS neutrophils, displayed as log2 fold change over HC, * P ≤ 0.05, ** P ≤ 0.01, *** P ≤ 0.001, **** P ≤ 0.0001; B-C: *BTHS5* – non-neutropenic BTHS patient. **D.** Western blot of selected UPR-related proteins in HC or BTHS patient HSC-derived neutrophils at the end of differentiation (D17), n = 3 (HC, BTHS); asterisks indicate nonspecific bands. **E.** Average percentage of HSC-derived neutrophils (CD66b^+^CD15^+^) and mature neutrophils (CD66b^+^CD15^+^CD11b^hi^) at D17, after treatment with PERK inhibitor (GSK2606414; 1 µM, added on day 3, 5, 7, 10, and 14 of culture) or vehicle control (DMSO), n = 9 (DMSO, PERKi); * P ≤ 0.05. **F.** Average percentage of HSC-derived neutrophils (CD66b^+^CD15^+^) and mature neutrophils (CD66b^+^CD15^+^CD11b^hi^) at D17, after treatment with PERK activator (CCT020312; 1 µM, added on day 3, 7, 10, and 14 of culture) or vehicle control (DMSO), n = 4 (DMSO, PERKa); ns – not significant.

To confirm these changes, we quantified UPR protein abundance by Western blot (Fig. 5D). To avoid confounding by G-CSF treatment, we used HSC-derived neutrophils from BTHS patients and HC, which are exposed to equivalent amounts of G-CSF over the 10 days of the differentiation protocol. Despite limited sample size, immunoblotting confirmed overexpression of IRE1α, BiP, HSP60, and enhanced phosphorylation of eIF2α (Fig. 5D, S5E).

UPR signaling is controlled by three upstream master regulators: ATF6, IRE1ɑ and PERK. We tested pharmacological inhibitors targeting all 3 of these pathways in differentiation of neutrophils from HSC and found that inhibition of UPR regulator PERK with GSK2606414 leads to a partial block in neutrophil differentiation, as evidenced by the reduced number of CD66b^+^CD15^+^ (DMSO = 51.12±15.79% vs. PERKi = 39.02±18.92%; p = 0.043) and CD11b^hi^ (DMSO = 39.43±14.19% vs. PERKi = 24.58±14.56%; p = 0.043) neutrophils (Fig. 5E), reduced CD11b surface expression (DMSO = 1.00±0.44 vs. PERKi = 0.77±0.38, p = 0.254); (Fig. S5F), and diminished proliferation by the end of the differentiation process (DMSO = 27±16 vs. PERKi = 9±8; p = 0.0064); (Fig. S5G). ATF6 inhibitor (Ceapin A7) and IRE1α inhibitor (4μ8C) did not affect development at the concentration tested (data not shown). To further investigate the role of the PERK pathway in neutrophil differentiation, we used the PERK activator CCT020312 to increase the activity of this signaling pathway. Similarly to PERK inhibition, PERK activation exhibited a trend towards partial reduction of neutrophil differentiation, as evidenced by reduction in numbers of CD66b^+^CD15^+^ cells (DMSO = 61.73±9.39% vs. PERKa = 53.30±5.54%; p = 0.185) and CD11b^hi^ cells (DMSO = 54.00±10.67% vs. PERKa = 43.00±7.95%; p = 0.153); (Fig. 5F), and reduced CD11b surface expression (DMSO = 1.00±0.18 vs. PERKa = 0.78±0.08, p = 0.067); (Fig. 5H) although neither were statistically significant. We also observed a similar trend towards diminished proliferation by the end of the differentiation process (DMSO = 41±11 vs. PERKa = 32±12; p = 0.301); (Fig. 5I), although again these differences were not significant. Taken together, these results implicate dysregulation of unfolded protein response in impairment of neutrophil development in BTHS.

## DISCUSSION

Despite the prominence of neutropenia in BTHS patients, our understanding of the molecular and physiological causes of this phenomenon has been limited. Our study reveals that tafazzin has a cell-intrinsic role in regulating neutrophil differentiation and confirms a partial block in granulopoiesis in BTHS patients. We also show that tafazzin deficient neutrophils exhibit enhanced degranulation and elevated rate of NETosis. Finally, we confirm alterations in UPR signaling in BTHS neutrophils, which might be a contributing factor to the observed developmental and functional changes.

We observed a decrease in the number of mature neutrophils in BTHS bone marrow samples. This is consistent with the original bone marrow examination by Barth [18] (n=2) and subsequent work [12] that demonstrated hypocellularity in some, but not all, BTHS patients. It remains unclear whether reduction in bone marrow neutrophils exists in a subset of patients and whether this is defined by the nature of the tafazzin mutation or by extrinsic factors. The maturation deficiency largely manifests in later stages of neutrophil development, affecting metamyelocytes and band neutrophils. How this leads to neutropenia, rather than simply a decrease in maturity of circulating neutrophils, remains unexplained, and implies the involvement of additional extrinsic factors.

We did not observe any evident hallmarks of early apoptosis in BTHS cultured neutrophils, which is reflected in largely unaltered proliferation rates. We showed that primary BTHS neutrophils exhibited a modest increase in annexin V binding, which is also induced by fMLP stimulation. Consistent with the conclusion of Kuijpers et al. [19] that increased binding of annexin V does not correlate with apoptosis, we propose that elevated PS in BTHS neutrophils results from increased neutrophil degranulation, which is known to alter surface lipid composition and transiently expose PS [31, 32]. We conclude that neutropenia in BTHS patients is most likely not caused by accelerated apoptosis.

Intriguingly, we also observed increased functional responses in tafazzin deficient neutrophils. Both spontaneous and bacterial-induced degranulation were elevated in patient neutrophils, although this was not observed when cells were cultured from tafazzin knockout stem cells. This discrepancy may be due to the fact that knockout neutrophils display a more severe developmental delay than patient cells and the propensity to hyperdegranulate is masked by developmental delays in synthesis of granule components in KO neutrophils. Alternatively, unknown circulating factors in patient plasma, such as inflammatory cytokines, may be priming circulating neutrophils and lowering their degranulation threshold.

Tafazzin knockout neutrophils also show elevated rates of inflammatory NETotic cell death in response to calcium mobilization, although this was not tested in circulating BTHS patient neutrophils. Increased NET formation may lead to peripheral depletion of neutrophils, possibly contributing to neutropenia. NET release is also a potent inflammatory signal that activates macrophages and other immune cells [33, 34], potentially lowering the threshold for acute and chronic inflammation [35, 36]. These findings highlight a major knowledge gap on inflammatory processes in BTHS and suggest that inflammation should be further investigated as an etiological factor in BTHS.

Several mechanisms can be proposed for enhanced granule exocytosis and NETosis in tafazzin deficient neutrophils. First, we observed elevated levels of intracellular calcium in BTHS cultured and primary neutrophils; Ca^2+^ flux promotes neutrophil degranulation [27, 29] and NET release [30]. Mitochondria are known to regulate intracellular calcium stores [37] and this mechanism may be perturbed by alterations in cardiolipin composition [10]. Secondly, it is known that ROS production, including mtROS, can trigger the disassembly of filamentous actin (F-actin) [38–40], which is needed for both mobilization of granules [41, 42] and NET formation [43]. We observed elevated mtROS in BTHS neutrophils, suggesting that activation in BTHS may be facilitated by ROS-induced actin degradation. Finally, degranulation is regulated by UPR sensors, which we found to be dysregulated in both primary and cultured BTHS neutrophils. In a model of acute lung injury, ER stress triggered by the IRE1/XBP1 pathway promotes degranulation, and specific depletion of XBP1 in neutrophils reduces granule secretion [44]. Similarly, in lupus, neutrophils exhibit elevated IRE1α activity that was correlated with increased extracellular elastase activity [45]. These findings imply that fluctuations in UPR during the physiological turnover of neutrophils could influence their inflammatory response.

In contrast to previous findings associating tafazzin deficiency with disruptions in mitochondrial respiration [46–49], we found no defects in the basal and maximal respiratory rates nor in the rate of ATP production in circulating BTHS neutrophils, although the interpretation of these findings is complicated by the fact that G-CSF-mobilized immature neutrophils have elevated mitochondrial content and activity [24]. Neutrophils differ from other cells in that they have fewer mitochondria per cell, which has traditionally led to the belief that these cells primarily rely on glycolysis and are not dependent on active mitochondria; however, recent advancements have highlighted the significance of these organelles in neutrophil development and function [50, 51]. Future studies should carefully examine mitochondrial metabolism in BTHS progenitors.

Interestingly, our proteome analysis uncovered alternations in UPR and mtUPR pathways in BTHS neutrophils, which is in accordance with findings from *Tafazzin*-KO mice [23]. UPR pathways were traditionally linked with protein misfolding in the ER; however, recently these signaling modules were recognized to play a crucial role in regulation of immunity and inflammation [52]. For instance, experiments using HL-60 cells demonstrated that *in vitro* neutrophil differentiation relies on the stage-specific expression of canonical UPR regulators. Notably, inhibition of these three proteins reduced the expression of CD11b and morphological differentiation, which we also confirmed in our primary cell culture system using PERK inhibitor GSK2606414. Although the studies using *Tafazzin*-KO mice and HL-60 cells linked perturbations in UPR with increased apoptosis [23, 53], as discussed above, we did not observe differences in cell survival in cultured and circulating BTHS neutrophils. Neutropenia is one of the symptoms of Wolcott-Rallison syndrome caused by mutations in the *EIF2AK3* gene, which encodes PERK [54], and heightened ER stress was reported in neutropenias caused by mutations in *ELANE* and glucose-6-phosphate subunit α gene (*G6PC3*); [55–57]. These findings collectively highlight the importance of tightly regulated UPR in neutrophil development.

In summary, we confirmed a partial block in neutrophil maturation in BTHS patients, resulting in generation of fewer mature neutrophils from stem cells. Despite alterations in phenotype and function, BTHS neutrophils maintain their anti-microbial activity and display a hyperinflammatory phenotype. Potential causes of these changes could be alterations in the UPR and calcium signaling pathways, which are currently recognized as vital for neutrophil development and function. As neutropenia in BTHS patients has irregular patterns, disruption of neutrophil differentiation may be triggered under non-homeostatic conditions such as metabolic perturbations or other forms of cellular stress. These conundrums imply the presence of underlying imbalances alongside functional neutrophil precursors, necessitating further research to understand the nuanced mechanisms at play.

## MATERIALS AND METHODS

### Human subjects and samples

The study was approved by NHS Research Ethics committee (permit number 09/H0202/52). Written informed consent was received from all patients and healthy donors. The patient cohort consisted of 28 patients (27 male), while the controls consisted of 31 individuals (27 male). Samples from some patients and healthy donors were obtained multiple times. Patient genetic data (*TAFAZZIN* mutations) are listed in Table 1. Venous blood was collected in EDTA tubes (BD Biosciences). For shRNA and CRISPR/Cas9 experiments, HSC were isolated from apheresis blood (NHSBT, Filton, Bristol, UK) with NHS REC approval (18/EE/0265).

**Table 1.** Genetic information of patients.

Genetic information of patients
| mutation | effect | exon/intron |
| --- | --- | --- |
| c.9_10dupG | p.His4Alafs*130; frameshift | exon 1 |
| c.51G>A | p.Trp17*; nonsense | exon 1 |
| c.82_84delGTG | p.Val28del; deletion | exon 1 |
| c.118A>G<sup>1</sup> | p.Asn40Asp; missense | exon 2 |
| c.149T>C | p.Leu50Pro; missense | exon 2 |
| c.182del | deletion/frameshift | exon 2 |
| c.207C>G | p.His69Gln; missense | exon 2 |
| c.216C>A | p.Cys72*; nonsense | exon 2 |
| c.228_232del | deletion/frameshift | exon 2 |
| c.239G>A | p.Gly80Glu or RNA splicing; missense | exon 3 |
| c.281G>A<sup>1</sup> | p.Arg94His; missense | exon 3 |
| c.346G>C | p.Gly116Arg; missense | exon 3 |
| c.547_548insdel | insertion-deletion | exon 7 |
| c.553A>G | new exonic splicing donor at codon Met185; p.Lys182Glnfs*4; frameshift | exon 7 |
| c.581G>A | p.Trp194*; nonsense | exon 7 |
| c.583+5G>A | GTAAGG>GTAAAG; RNA splicing | intron 7 |
| c.589G>A<sup>2</sup> | p.Gly197Arg; missense | exon 8 |
| c.589G>T | p.Gly197Trp; missense | exon 8 |
| c.646+1del | deletion/frameshift | exon 8 |
| c.809_812del | deletion/frameshift | exon 11 |
| c.837_838delTC | p.Gln280Glyfs*30; frameshift | exon 11 |

<sup>1</sup>Variant present in 2 patients
<sup>2</sup> Variant present in 4 patients
For 1 patient variant was unknown.
For more details on *TAFAZZIN* variants please refer to Human *TAFAZZIN* Gene Variant Database at: https://www.barthsyndrome.org/research/tafazzindatabase.html

### Mouse model and experiments

All mouse experiments were performed in accordance with UK Home Office regulations (Project License PP9886217), under the oversight of the Animal Welfare and Ethical Review Board (AWERB) of the University of Glasgow.

The *Tafazzin* knockout mice were generated using G4 embryonic stem cells (isolated from C57BL/6Ncr x 129S6/SvEvTac F1 mice). After germline transmission of the targeted allele mice were bred for at least 10 generation to FVB/NCrl mice.

### CD34^+^ HSC isolation and neutrophil culture

CD34^+^ HSCs were isolated either from peripheral blood of consented healthy donors and BTHS patients or from apheresis blood and then cultured according to a modified protocol of Naveh et al. [24]. In brief, peripheral blood mononuclear cells (PMBCs) were isolated by density centrifugation using Histopaque®-1077 (Sigma-Aldrich) according to manufacturer’s instructions, followed by red cell lysis (55mM NH_4_Cl, 0.137mM EDTA, 1mM KHCO_3_, pH 7.5). CD34^+^ cells were enriched with a human CD34 Microbead Kit (Miltenyi Biotec) according to manufacturer’s protocol. For peripheral blood HSCs, the cells were cultured in StemSpan^TM^ Hematopoietic Cell Media (STEMCELL^TM^ Technologies) supplemented with 1% (v/v) penicillin-streptomycin (P/S, Sigma-Aldrich) for the first 4 days and from day 5 of the culture, cells were cultured in Iscove’s Modified Dulbecco’s Media (IMDM, Gibco^TM^) supplemented with 1% P/S and 10% (v/v) heat-inactivated fetal bovine serum (FBS, Sigma-Aldrich). Cells isolated from apheresis blood were cultured in IMDM only. Cytokines were added at the indicated concentrations and days of culture: stem cell factor (SCF, 50 ng/mL; day 0-5 of culture), Flt-3 ligand (50 ng/mL; day 0-5 of culture), interleukin-3 (IL-3, 10 ng/mL; day 0-5 of culture), granulocyte-macrophage colony-stimulating factor (GM-CSF, 10 ng/mL; day 3-7 of culture), and granulocyte colony-stimulating factor (G-CSF, 10 ng/mL; day 7-14 of culture). All inhibitors or activators were added on day 3, 5, 7, 10 and 14 of the culture. All functional assays (unless stated otherwise) were completed between Day 17 and 19 of culture.

### CRISPR-mediated knockout of *TAFAZZIN*

CRISPR/Cas9 genome editing was performed on day 3 cultured neutrophils, by nucleofection of ribonuclear particles, as described in [24]. gRNAs were designed using Knockout Guide Design (Synthego). The following sgRNAs were used (Synthego, modified sgRNA with EZ scaffold):

TAZ+154413511: UGCAGACAUCUGCUUCACCA;

TAZ-154413490: GCAGAUGUCUGCAGCUGCAG;

scrambled control #1: GCACUACCAGAGCUAACUCA;

scrambled control #2: GUACGUCGGUAUAACUCCUC.

### shRNA-mediated knockdown of *TAFAZZIN*

shRNA knockdown was achieved using lentiviral transduction of day 3 cultured HSC. HEK293T cells were transfected with psPAX2 and pMD2.G packaging vectors, along with U6-based shRNA lentiviral vectors, with enhanced green fluorescent protein (eGFP) as a marker (VectorBuilder Inc.). Virus was concentrated using Lenti-X^TM^ Concentrator and added to the cultured neutrophils.

shRNA sequences:

shRNA 1: TGCTTCCTCAGTTACACAAAGCTCGAGCTTTGTGTAACTGAGGAAGCA

shRNA 2:

CTGTGGCATGTCGGAATGAATCTCGAGATTCATTCCGACATGCCACAG

shRNA 3: CGGACTTCATTCAAGAGGAATCTCGAGATTCCTCTTGAATGAAGTCCG

n-t shRNA (scrambled control):

CCTAAGGTTAAGTCGCCCTCGCTCGAGCGAGGGCGACTTAACCTTAGG

### Flow cytometry analysis

The analysis was conducted as described in [25]. Briefly, cells were stained with 0.1% Zombie^TM^ live/dead stain (BioLegend) and incubated for 10 min in the dark at RT. Samples were washed, incubated in Fc receptor blocking solution Human TruStain FcX^TM^ (BioLegend) diluted in MACS buffer (5 mM EDTA, 0.5% BSA in PBS) for 5 min, and then a mix of primary antibodies in MACS was added followed by 30 min incubation on ice. Samples then were washed twice with MACS buffer and fixed for 20 min with 2% formaldehyde in PBS. Cells were analyzed with BD LSRFortessa™ X-20 Cell Analyzer. OneComp eBeads^TM^ (Invitrogen^TM^) were used as a single-color compensation control. For Annexin V positivity, cells were stained with FITC-Annexin V (BioLegend). At least 10,000 events were recorded per sample, and then analyzed in FlowJo^TM^ software (version 9).

### Western blot

Cells were lysed directly in 2x Bolt^TM^ LDS sample buffer (Invitrogen^TM^) supplemented with Halt^TM^ Protease and Phosphatase Inhibitor Cocktail (Thermo Scientific^TM^) and heated for 10 min at 70°C. Samples were sonicated and then run on 4-12% Bolt^TM^ Bis-Tris Plus Mini Protein gels (Invitrogen^TM^), followed by transfer onto Immobilon^®^-FL PVDF membrane (MERCK), using a standard wet transfer protocol. The membrane was blocked with 5% milk, incubated with primary antibodies overnight, and on the following day with secondary antibodies for 1 h at RT. Blots were imaged using a LI-COR Odyssey^®^ XF Imager.

### PLB-985 cell culture and differentiation

PLB-985 were cultured in RPMI-1640 (Sigma-Aldrich) supplemented with P/S and 10% FBS. After lentiviral transduction, cells were differentiated by replacing the media with RPMI-1640 with P/S, 2.5% FBS, 0.5% DMF, 1x Nutridoma-CS (Roche) and culturing for 7 days.

### Neutrophil isolation, oxidative burst, and NET formation

Neutrophils were isolated using the EasySep^TM^ Direct Human Neutrophil Isolation Kit (STEMCELL^TM^ Technologies) as per the manufacturer’s instructions. ROS production was quantified with the luminol method, as previously described [58]. Chemiluminescence was recorded in 2-minute intervals using a FLUOstar^®^ Omega plate reader (BMG LABTECH). NET assays were performed as described in [59]. Cells were stimulated with 50nM PMA, 300nM fMLP, or 10μM A23187. NETs were stained with SYTOX and SYTO dyes and imaged with either an EVOS® FL or Incucyte® ZOOM imaging systems.

### MPO ELISA

Plasma MPO was quantified using Human Myeloperoxidase DuoSet ELISA (R&D Systems), according to manufacturer’s instructions.

### Degranulation

In experiments on circulating neutrophils, cells were stimulated with N-formylmethionine-leucyl-phenylalanine (fMLP, 300nM) for 30 min or *Streptococcus pyogenes* MGAS10270 for 40 min. For experiments on HSC-derived neutrophils, cells were stimulated with A23187 (2.5μM) for 30 min. After stimulation, cells were stained and fixed according to the flow cytometry protocol detailed above.

### Bacterial killing assay

Neutrophils (2.5×10^6^) were resuspended in 500μL HBSS and combined with 5×10^5^ *Streptococcus pyogenes* MGAS10270 bacteria [60] in HBSS supplemented with 2mM CaCl_2_, 2mM MgCl_2_ and 10% pooled human serum (SEQUENS IVD). Samples were incubated at 37°C with atmospheric CO_2_ levels on a rotator. Aliquots (50μL) of each sample were taken immediately and thereafter every hour for 4 hours, and numbers of viable bacteria enumerated following plating onto Todd-Hewitt agar plates supplemented with 0.5% (w/v) yeast extract and incubation overnight at 37°C, 5% CO_2_. Killing efficiency was calculated as number of colonies with neutrophils/number of colonies with serum only.

### Fluorescent immunohistochemistry and mouse whole blood analysis

Lung and spleen sections of germline *Tafazzin* −/− mice, were stained for neutrophil abundance with a neutrophil-specific calgranulin antibody, as previously described [61]. Tissue sections were imaged with a Leica DMI6000 inverted epifluorescence microscope and analysis was performed with ImageJ Fiji. For whole blood analysis, mice were tail vein bled, and blood sample analysis was performed using the Procyte DX Hematology analyzer.

### Phagocytosis

Heat-killed *Streptococcus pyogenes* MGAS10270 (1×10^9^) were stained with pHrodo^TM^ Phagocytosis Particle Labeling Kit for Flow Cytometry (Invitrogen^TM^) following the manufacturer’s protocol. Neutrophils (2×10^6^) were resuspended in 500 μL RPMI+Q and 2×10^8^ pHrodo-stained *Streptococcus pyogenes* were added for an MOI = 100. Suspensions were incubated with rotation at 37°C for 1 h, taking 100 μl aliquots at multiple time points. Cells were then washed once and analyzed using BD LSRFortessa™ X-20 Cell Analyzer.

### Seahorse metabolic flux analysis

Seahorse assay was performed according to the protocol in [24] using Seahorse XFe96 microplates (Agilent) and Seahorse XF DMEM medium (Agilent).

### Mitochondrial labelling

To label mitochondria, cells were incubated with 25 nM tetramethylrhodamine, ethyl ester, perchlorate (TMRE, Invitrogen^TM^), 5 µM MitoSox^TM^ Red (Invitrogen^TM^), or 5 nM MitoTracker^TM^ green (Invitrogen^TM^) in prewarmed HBSS. Cells were incubated with each dye for 20 min at 37°C, washed with fresh media, resuspended in MACS buffer, and analyzed using BD LSRFortessa™ X-20 Cell Analyzer.

### Intracellular calcium

Levels of intracellular Ca^2+^ were determined using the cell permeant calcium indicator X-Rhod-1, AM (Invitrogen^TM^). Cells were incubated with 500 nM X-Rhod-1 in RPMI for 1 h at 37°C, thoroughly washed with fresh media, resuspended in MACS buffer, and analyzed with BD LSRFortessa™ X-20 Cell Analyzer.

### Proteomics and bioinformatics analysis

Isolated peripheral blood neutrophils from patients and controls were lysed and labeled with tandem mass spectrometry reagents (Thermo Fisher), as previously described [25]. Peptides were identified by nano LC-MS/MS with a Orbitrap^TM^ Fusion Tribrid^TM^ Mass Spectrometer (Thermo Scientific^TM^). Raw files were analyzed using Proteome Discoverer^TM^ software v. 2 and cross-referenced against the human UniProt database (human). The protein groups were reassessed by an in-house script which initially selects a master protein by ID and quantitation metrics, then by annotation quality of Uniprot accessions. Data were log_2_ transformed and tested for statistical significance using Welch’s *t* test. IPA analysis was performed with a filter of P < 0.05 to identify biological trends in the proteins that were statistically significant between conditions. PCAs were calculated using the PCA function in the FactoMineR package and plotted using either ggplot (2D) or Plotly (3D).

### Statistical analysis

The analysis was performed using GraphPad Prism 8 software. All the data are presented as mean ± SD. Unless stated otherwise, all “n” numbers represent the number of blood samples. Statistical analysis was completed, where appropriate, using t test when comparing two different samples, or one-way/two-way ANOVA where multiple samples were being examined.

## FIGURE LEGENDS

**Fig. S1.**
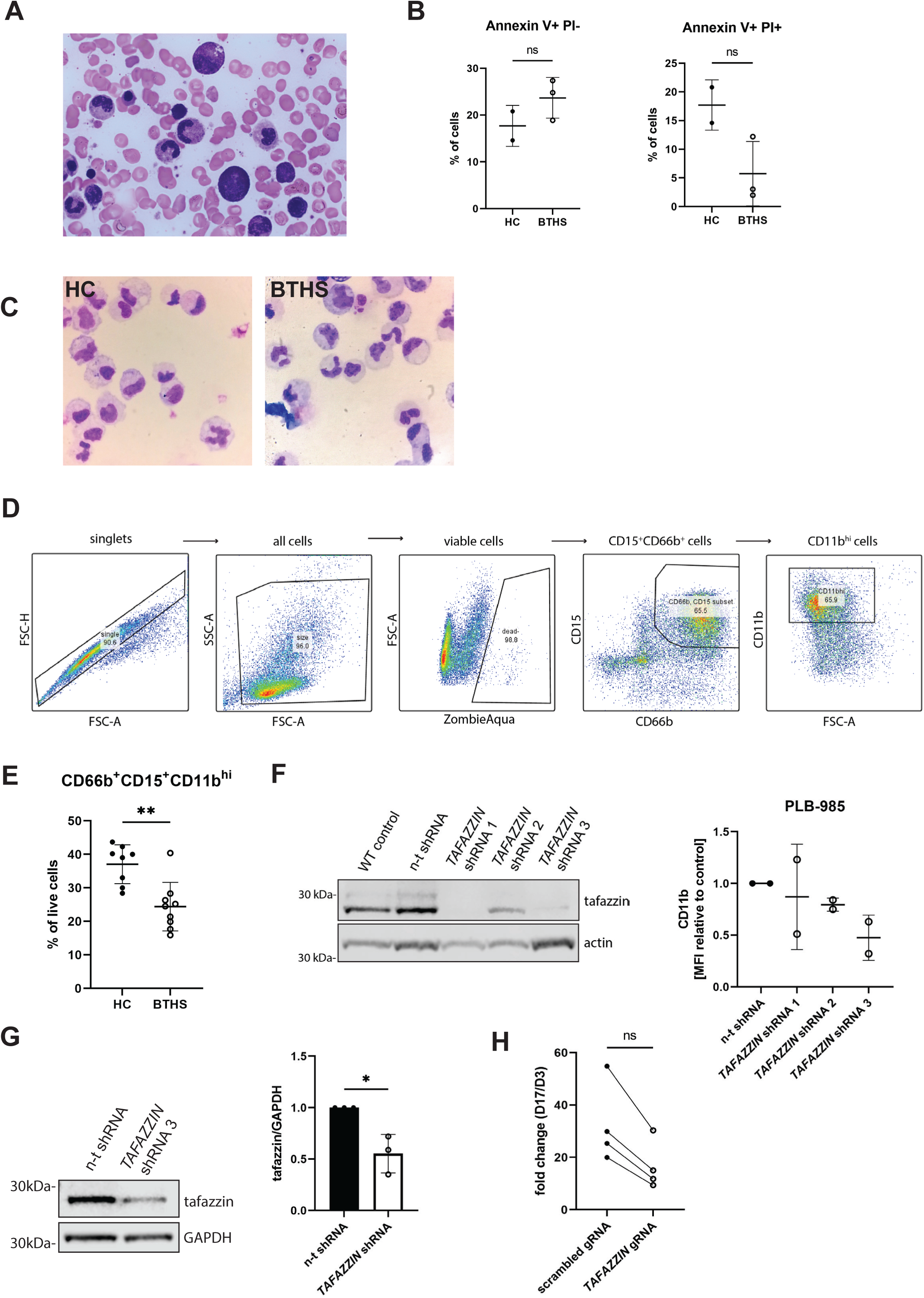
Impaired terminal differentiation and maturation of BTHS neutrophils *ex vivo*. **A.** Representative image of BTHS patient bone marrow aspirate. **B.** Quantification of average percentage of apoptotic (AV+PI-) and necrotic (AV+PI+) HSC-derived neutrophils at the end of differentiation, n = 2 (HC), 3 (BTHS); ns – not significant. **C.** Representative cytospins of HSC-derived neutrophils (day 17). **D.** Gating strategy for HSC-derived neutrophils (day 17). **E.** Average percentage of CD66b^+^CD15^+^CD11^hi^ cells in live population of HSC-derived neutrophils at day 17, n = 9 (HC), 8 (BTHS) **F.** Representative Western blot depicting level of tafazzin expression and graph showing CD11b surface expression of PLB-985 cells transduced with lentivirus encoding non-targeting (n-t) or anti-*TAFAZZIN* shRNAs, n = 2 (experimental repeats). **G.** Representative Western blot and graph depicting level of tafazzin expression level of GFP^+^ HSC-derived neutrophils transduced with lentivirus encoding non-targeting (n-t) or anti-*TAFAZZIN* shRNAs, n = 3 (experimental repeats), * P ≤ 0.05. **H.** Fold change of total cell count of CRISPR/Cas9-edited HSC-derived neutrophils from day 3 to day 17 of differentiation matching tafazzin knockout cells to their relative controls, n = 4; * P ≤ 0.05.

**Fig. S2.**
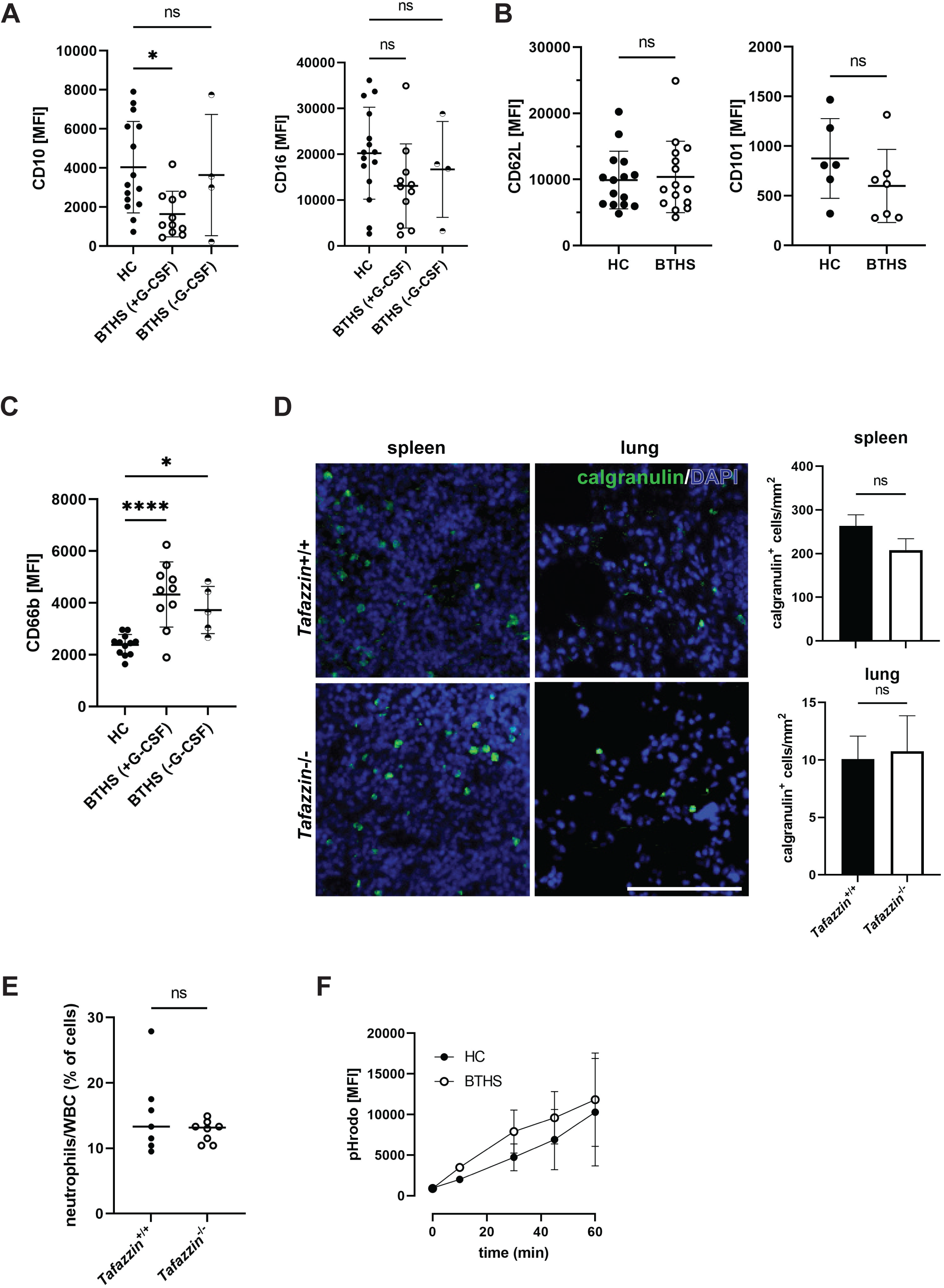
Additional analyses of peripheral blood neutrophils. **A.** Surface expression of CD10 (left) and CD16 (right) in circulating neutrophils stratified according to G-CSF therapy; n = 15 (HC), 11 (BTHS+G-CSF), 4 (BTHS−G-CSF); * P ≤ 0.05, ns – not significant. **B.** Surface expression of CD62L (left) and CD101 (right) in circulating neutrophils, n = 15 (HC, BTHS); ns – not significant. **C.** Surface expression of CD66b in neutrophils stratified according to G-CSF treatment, n = 12 (HC), 10 (BTHS+G-CSF), 5 (BTHS−G-CSF); * P ≤ 0.05, **** P ≤ 0.0001. **D.** Representative epifluorescence images showing anti-calgranulin (green) and DAPI (blue) staining of mouse lung and spleen (left) and quantification of average number of calgranulin-positive cells per mm^2^ of section (right), n = 2 (mouse per genotype; the cells were counted from one transverse section through the middle part of the tissue). **E.** Percentage of neutrophils in mouse whole blood, n = 7 (WT), 8 (KO), ns – not significant. **F.** Phagocytosis time-course, measured by pHrodo fluorescence in isolated neutrophils, n = 3 (HC, BTHS).

**Fig. S3.**
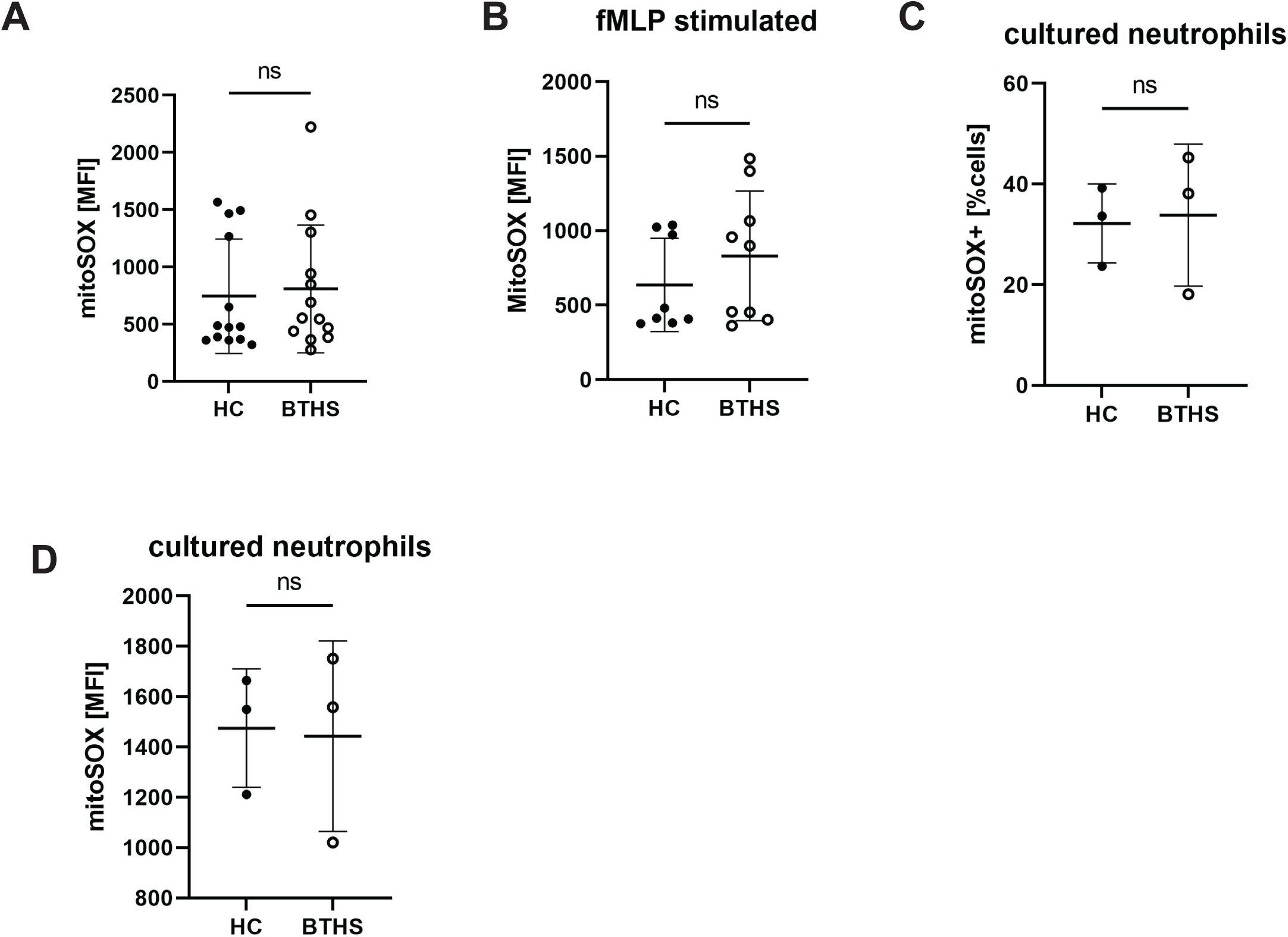
Additional mitochondrial analyses in BTHS neutrophils. **A.** Quantification of average mitoSOX median fluorescence of circulating neutrophils, n = 13 (HC, BTHS); ns – not significant. **B.** Quantification of average mitoSOX median fluorescence of circulating neutrophils stimulated with 300 nM fMLP, n = 8 (HC), 9 (BTHS), ns – not significant. **C.** Percentage of mitoSOX-positive HSC-derived neutrophils at the end of differentiation (D17), n = 3 (HC, BTHS); ns – not significant. **D.** Percentage of mitoSOX-positive HSC-derived neutrophils (D17), after stimulation with 300 nM fMLP, n = 3 (HC, BTHS); ns – not significant.

**Fig. S5.**
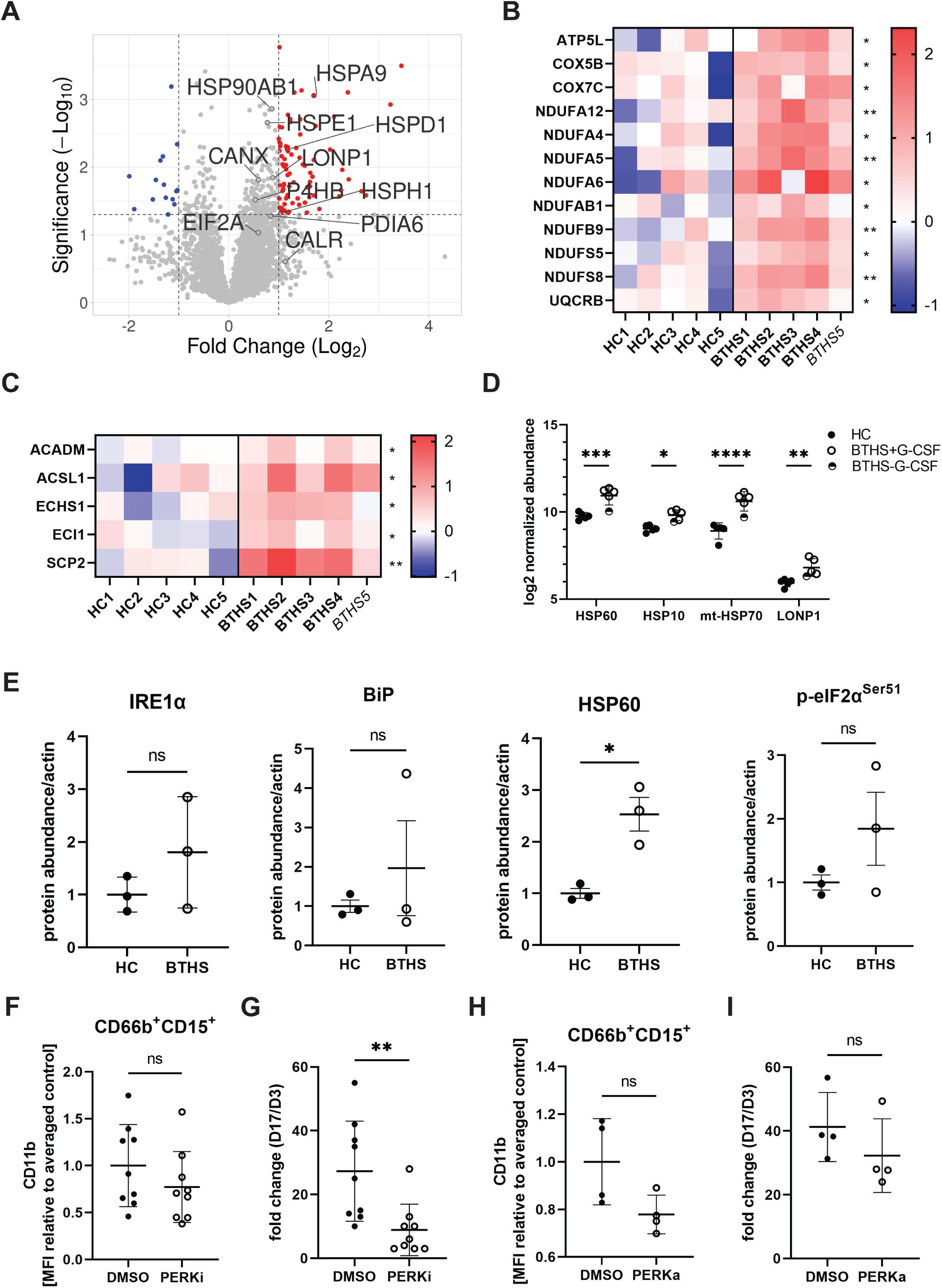
Elevated UPR signaling in BTHS neutrophils. **A.** Volcano plot comparing BTHS and HC circulating neutrophil protein abundances, displayed as −log_10_ *P*-value and log_2_ fold change. **B.** Heat map depicting upregulated oxidative phosphorylation proteins identified with IPA, displayed as log2 fold change over HC, * P ≤ 0.05, ** P ≤ 0.01. **C.** Heat map depicting upregulated fatty acid oxidation proteins identified with IPA, displayed as log2 fold change over HC, * P ≤ 0.05, ** P ≤ 0.01; B-C: *BTHS5* – non-neutropenic BTHS patient. **D.** Log_2_ normalized abundance of mtUPR-related proteins identified by proteomics in circulating neutrophils, * P ≤ 0.05, ** P ≤ 0.01, *** P ≤ 0.001, **** P ≤ 0.0001. **E.** Quantification of UPR-related protein expression in HSC-derived neutrophils at D17 of differentiation, n = 3 (HC, BTHS); ns – not significant, * P ≤ 0.05. **F.** Surface expression of CD11b in PERK-inhibited cells (GSK2606414; 1 µM) or vehicle control (DMSO-treated) HSC-derived neutrophils at D17 of differentiation, relative to averaged control (on a same day), n = 9 (DMSO, PERKi); ns – not significant. **G.** Fold change in total cell count during HSC differentiation, from day 3 to day 17, after treatment with PERK inhibitor (GSK2606414; 1μM) or vehicle control (DMSO), relative to averaged CD11b expression of vehicle control cells on a same day, n = 9 (DMSO, PERKi); ** P ≤ 0.01. **H.** Surface expression of CD11b in PERK-activated cells (CCT020312; 1 µM) or vehicle control (DMSO-treated) HSC-derived neutrophils at D17 of differentiation, relative to averaged control (on a same day), n = 4 (DMSO, PERKa); ns – not significant. **I.** Fold change in total cell count during HSC differentiation, from day 3 to day 17, after treatment with PERK activator (CCT020312; 1 µM) or vehicle control (DMSO), relative to averaged CD11b expression of vehicle control cells on a same day, n = 4 (DMSO, PERKa); ns – no significant.

## DECLARATIONS

### Ethics approval and consent to participate

Patient blood samples were collected with approval from National Health Service Research Ethics Committee (REC), 09/H0202/52. Apheresis and healthy control blood samples were collected under NHS REC 18/EE/0265.

### Availability of data and materials

The research materials supporting this publication can be accessed by contacting corresponding authors.

The mass spectrometry proteomics data have been deposited to the ProteomeXchange Consortium via the PRIDE [62] partner repository with the dataset identifier PXD052714.

### Author contributions

**PZ, CR, DC, SG, FP, WG, KR, TP, EA** and **BA** performed experiments and analyzed data; **KF** organized access to and analysis of bone marrow aspirates; **DS** provided mouse samples; **AHN** supervised bacterial experiments; **BA, CS and AT** conceived and supervised the study and acquired funding. **PZ** and **BA** wrote the manuscript.

All authors read, provided input, and approved the final manuscript.

## Data Availability

All data produced in the present study are available upon reasonable request to the authors.

## ACKNOWLEDGMENTS

We thank all the patients and blood donors for participating in our study, as well as nurses and clinical staff at the NHS Barth Syndrome Service and Bristol Children’s Hospital. We acknowledge the assistance and support from Gillian Alexander, Maria Pelidis, Effie Chronopoulou, Rachel Schwartz, Olivia Gordon, Sally Turner, and Germaine Pierre. We also thank Jane Brittan for technical assistance with the bacterial experiments.

